# Effects of statin therapy on coronary plaque volume by decreasing CRP/hsCRP levels: A meta-regression of randomized controlled trials

**DOI:** 10.1101/2021.12.26.21268412

**Authors:** Darui Gao, Rong Hua, Dina Jiesisibieke, Yanjun Ma, Chenglong Li, Sijing Wu, Qian Ma, Wuxiang Xie

## Abstract

**Background and aims:** Several clinical trials have indicated that statins stabilize and reverse atherosclerotic plaque. However, different studies have provided inconsistent findings regarding mechanisms and influencing factors of plaque regression under statin therapy. In this study, meta-analysis and meta-regression were used to determine the effects of statin medications on coronary plaque volume as determined by intravenous ultrasound. Meanwhile, the impact of CRP/hsCRP reduction during statin therapy on plaque regression was investigated.

**Methods:** Up to May 28, 2021, a systematic PubMed, EMBASE, and Cochrane search was performed for randomized controlled trials that assessed treatment effect using total atheroma volume (TAV), percent atheroma volume (PAV), or plaque volume (PV). Only CRP/hsCRP and LDL-C values reported before and after treatment were considered.

**Results:** 12 studies fulfilled the inclusion criteria and were included in the systematic review. A meta-analysis of 15 statin-treated arms reported a significant reduction in change of TAV/PV (standardized mean difference [SMD]: −0.27, 95% confidence intervals [CI]: −0.42, −0.12), compared with the control arms. Another meta-analysis of 7 trials also found that patients in the intervention group had a significant reduction in change of PAV (SMD: −0.16, 95% CI: −0.29, −0.03), compared with those in the control group. Meta-regression analysis revealed the percent change of CRP/hsCRP statistically influences SMD in change of TAV/PV after adjusting for percent change of LDL-C, age and gender. Meta-regression analysis showed that percent change of CRP/hsCRP statistically influences SMD in change of PAV.

**Conclusion:** In conclusion, statin therapy is beneficial for plaque regression. Statins promote plaque regression through their anti-inflammatory ability.

## Introduction

Cardiovascular diseases are considered the leading causes of death worldwide. Among them, coronary heart disease (CHD) has garnered considerable attention due to its high prevalence and burden. The pathological basis of CHD is atherosclerosis, which is characterized by accumulation of lipid and cholesterol in the artery’s subintima and progressive chronic inflammation of the fibrotic plaque on the wall of great and medium arteries [1].

Coronary plaque regression has a significant positive correlation with low density lipoprotein cholesterol (LDL-C). As important lipid-lowering drugs, several studies have demonstrated that statin drugs promote coronary atheroma stabilization and regression in patients with acute coronary events or stable coronary disease [2]. Currently, statin has become an important preventive drug for atherosclerotic cardiovascular disease (ASCVD). Numerous studies showing that statins are effective in reducing low-density lipoprotein cholesterol levels, and the risk of death and recurrent coronary and cardiovascular events in those with a history of ASCVD [3]. As the mechanism of vascular inflammation is gradually elucidated, numerous evidences demonstrate that C-reactive protein (CRP) and high-sensitivity C-reactive protein (hsCRP) may play direct pathogenic roles in atherosclerosis [4, 5]. Initially, statin drugs were used primarily to reduce blood lipids. With the deepening of research, its non-lipid-lowering effects, such as anti-inflammatory effect of statin on the coronary plaque volume has become the focus of recent studies. Ridker et al. discovered that rosuvastatin (20 mg/d) and placebo were administered to randomly selected healthy people with elevated hs-CRP but no evidence of hyperlipidemia. After an average follow-up of 1.9 years, hs-CRP level in the treatment group decreased by 37% compared with the control group, implying that statins may have anti-atherosclerosis functions via anti-inflammatory mechanisms [6]. Numerous clinical trials, such as the Air Force/Texas Coronary Atherosclerosis Prevention (AFCAPS/TexCAPS) study, the Reversal of Atherosclerosis with Aggressive Lipid Lowering (REVERSAL) trial, and the Pravastatin or Atorvastatin Evaluation and Infection Therapy-Thrombolysis In Myocardial Infarction 22 (PROVE IT-TIMI 22) trial, have demonstrated that statins reduced hsCRP levels independently of lowering LDL-C levels. In a trial with canakinumab for atherosclerotic disease, the rate of cardiovascular event recurrence was significantly lower in the treated group than in the placebo group, implying that reducing inflammation without affecting lipid levels can reduce cardiovascular disease risk [7].

Statin therapy was shown to be beneficial in reducing CRP/hsCRP. However, no study has attempted to investigate the relationship between the degree of CRP/hsCRP reduction associated with changes in coronary plaque volume during the statin treatment. To answer the question whether the CRP/hsCRP lowering effect of statins could delay or reverse the progression of atherosclerosis, we conducted this study. The aim of the present study was to provide a systematic review and meta-regression analysis to examine the impact of statins on CRP/hsCRP reduction on coronary plaque volume. At the same time, we analyzed the joint effects of LDL-C and CRP/hsCRP changes on plaques.

## Methods

This work followed the Preferred Reporting Items for Systematic reviews and Meta-Analyses (PRISMA) and amendment to the Quality of Reporting of Meta-analyses (QUOROM) statement [8, 9].

### Search strategy and study selection

For this meta-analysis, we have conducted a search in PubMed, EMBASE and Cochrane Library to identify studies relevant to this topic from their beginning to May 28, 2021. The study selection was performed independently by 2-group investigators (CLL, YJM as group 1, and RH, DJ as group 2) using highly sensitive strategy.

Disagreements were resolved by consensus with a senior author (WXX). Here we shown the search strategy of PubMed: “((statin) OR (hydroxy-methyl-glutaryl-CoA) OR (HMG-COA) OR (pravastatin) OR (lovastatin) OR (simvastatin) OR (Atorvastatin) OR (fluvastatin) OR (Rosuvastatin) OR (Pitavastatin)) AND ((intravascular ultrasound) OR (IVUS) OR (plaque) OR (atheroma)) AND ((intravascular ultrasound) OR (IVUS) OR (coronary)) AND (Clinical Trial[ptyp]).” Supplementary 1 shows details of search syntax.

### Selection criteria

Studies were included according to the following criteria: (a) randomized controlled trials (RCTs); (b) investigating the impact of statin therapy on plaque volume using IVUS; (c) reporting at least one of the following data: total atheroma volume (TAV), plaque volume (PV) and percent atheroma volume (PAV); (d) with a follow-up longer than or equal to six months; (e) reporting LDL-C at baseline and the end of the study or reporting data of percent change of LDL-C; (d) reporting CRP or hsCRP before and after statin treatment (or percent change of CRP/hs-CRP);

Exclusion criteria included the following: (a) duplicate publication or secondary analyses of the same study population; (b) lack of sufficient information on baseline or follow-up IVUS data, LDL-C data and CRP/hsCRP data.

### Data extraction quality appraisal

The data were extracted from each study using standard tables. The extracted data included the following: study characteristics (the first author, title, publication time, number of patients, country, and study duration), patient characteristics (age and sex), intervention, control, method characteristics (randomization, blind implementation, and follow-up loss), and patient outcomes. For patient outcomes, we extracted TAV, PAV, or PV data as measured using IVUS technique, LDL-C data, CRP, hsCRP data (including values at baseline and endpoint) and other useful information.

After data extraction, we conducted statistical processing to calculate change of TAV, change of PV, change of PAV, percent change of LDL-C, percent change of CRP and percent change of hsCRP. Articles reported mean values and standard deviation (SD) of change of TAV/PV/PAV, the original number was entered. Some studies [10–13] did not report SD values, which were filled by using the SD of the baseline data of control group. 1 study [14] provided SE rather than SD, and then SD value was calculated based on SE value. IVUS efficacy endpoints were reported as medians, with distribution-free 95% confidence intervals (CI) in 2 articles (Nicholls, 2011, Nissen, 2004), the median reported in the original text was extracted, and SD was calculated by formula.

In terms of LDL-C, if the article reported percent change of LDL-C, the original number was entered; otherwise, percent change of LDL-C was calculated using the following formula:

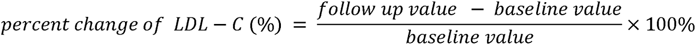

Percent change of CRP and percent change of hsCRP were calculated using the same approach. Supplementary 2 shows details of data extraction.

According to Cochrane’s indications, un-blinded, independent reviewers evaluated the quality of included studies using pre-specified forms (risk of bias table), including seven examined fields: random sequence generation (selection bias); allocation sequence concealment (selection bias); blinding of participants and personnel (performance bias); blinding of outcome assessment (detection bias); incomplete outcome data (attrition bias); selective outcome reporting (reporting bias); and other potential sources of bias.

### Data analysis and synthesis

Continuous variables were expressed as mean±SD, whereas categorical variables were expressed as *n* (%). Heterogeneity was evaluated by the *I^2^* test. A random-effects model was utilized. Meta-analysis with continuous outcome variables was performed, and the effect of statin therapy (vs. control) on change of TAV, PV and PAV at the end of follow-up was estimated as standardized mean difference (SMD) and 95% CI. If *p*<0.05 and 95% CI did not include zero, the point estimate of SMD was considered statistically significant. To avoid double-counting of subjects and consequent unit-of-analysis error in trials with more than one treatment arm, the control group was evenly divided (where possible) [10]. Since the units (mm^3^) of change of TAV and change of PV were the same, we combined these two indicators for data synthesis.

To explore the link between the dependent variable and the covariate, meta-regression is often used. We hypothesized that the included studies may have shown difference according to the percent change of CRP/hsCRP, percent change of LDL-C, age and gender of the patients. To evaluate the possible impact of these factors on the results of the meta-analysis, we established model with the change of TAV/PV or change of PAV as the dependent variable. In particular, change in TAV/PV was our primary outcome, and change in PAV was the secondary outcome.

Funnel plot analysis and Begg’s and Egger’s tests were performed to evaluate potential publication bias. Sensitivity analysis was conducted to assess the stability of studies. Sensitivity analysis was conducted using leave-one-out method, i.e. removing one study each time and repeating the analysis. Statistical analyses were performed with *R* version 4.1.2 (2021-11-01) and risk of bias was evaluated with Review Manager (RevMan 5.3; Cochrane Collaboration).

## Result

### Flowchart of included studies

The initial literature search retrieved 1259 articles. After the removal of duplicates, the titles and abstracts of 765 articles were carefully checked, leading to the exclusion of 626 articles for failing to meet inclusion criteria. Initially, 139 articles were selected, and their full texts were evaluated. Of them, 124 articles were excluded: 22 because CRP/hsCRP levels were not reported, 12 because plaque evaluation (TAV, PAV, or PV) was not performed, 50 because they were not RCTs, 31 because statins were not used, and 9 because of repeated trials. A total of 15 articles entered the third round of evaluation. One was excluded due to a discrepancy between the number of participants receiving statins and the number of people participating in IVUS measurements [15]. And two were excluded because of data quality: in one study CRP was reported in supplementary, but the indicators of the control group declined significantly [16]; in another study, the SD at baseline and follow-up varied greatly and the reported difference value was inconsistent with the calculated difference value [17]. Overall, this analysis included 12 trials [10–14, 18–24]. Figure 1 summarizes the study selection process.

**Figure 1.**
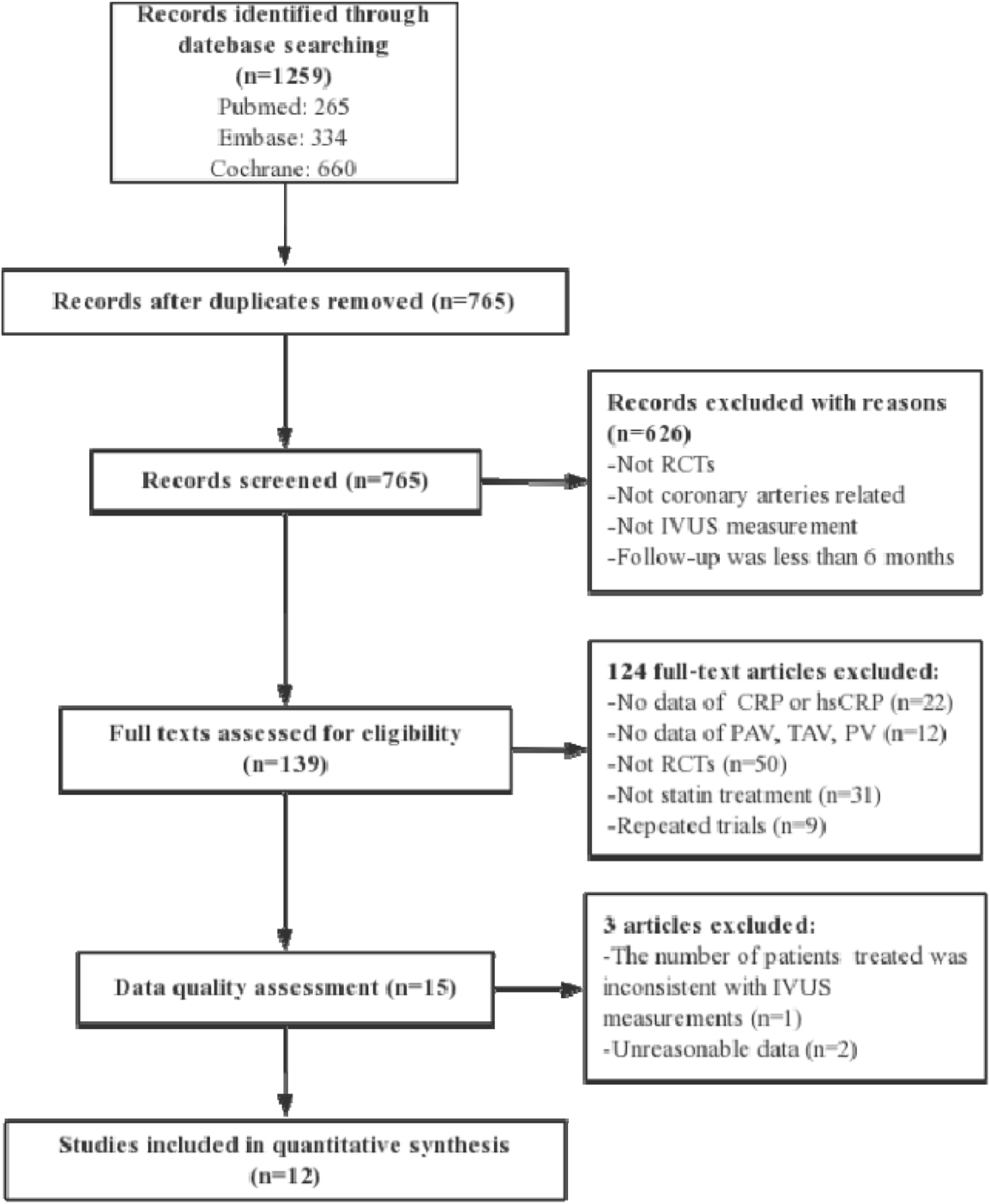
Flowchart for study. RCTs indicates randomized controlled trials; IVUS indicates intravenous ultrasound; CRP indicates C-reactive protein and hsCRP indicates high-sensitivity C-reactive protein. TAV indicates total atheroma volume; PAV indicates percent atheroma volume and PV indicates plaque volume.

### Characteristics of included studies

The study characteristics are reported in Table 1. A total of 2812 subjects were included in the twelve eligible studies. Included studies were published between 2004 and 2016 and were reported from China, American, Korea and Japan. The largest study had a population size of 1039 subjects while the smallest study recruited 30 subjects. The mean age of the participants ranged from 55.8 to 67.0 years.

12 trials with 16 treatment arms were included. 8 treatment arms used atorvastatin (dose range: 10-80 mg/day; duration of treatment: 24-72 weeks), 6 treatment arms used rosuvastatin (dose range: 10-40 mg/day; duration of treatment: 44-104 weeks), 1 treatment arm used pravastatin (dose: 20 mg/day; duration of treatment: 24 weeks), 1 treatment arm used pitavastatin (dose: 4 mg/day; duration of treatment: 32 weeks).

IVUS was used in all studies to evaluate plaque volume. In addition to 1 study [24], 11 studies reported change of TAV/PV, and 7 studies reported change of PAV. As described in the data extraction section, percent change of CRP/hsCRP and percent change of LDL-C were reported in all studies.

Overall, random sequence generation was observed in 6 studies, 4 of them reported allocation concealment. 3 trials were double-blinded, and 8 studies performed blinded assessments of the outcomes. Moreover, 2 studies existed incomplete outcome data because of high attrition rate. Supplementary 3 shows details of risk of bias assessment.

### Effect of statin therapy on change of TAV/PV

11 trials (n=2696) including 15 comparisons reported change of TAV/PV. Compared with control arms, our meta-analysis showed that 15 treatment arms revealed a significant decrease in change of TAV/PV (SMD: −0.27, 95% CI: −0.42, −0.12, *p*<0.05), with a moderate heterogeneity (*Q*=27.55, *df*=17, *p*=0.02, *I^2^*=49.2%). Figure 2 presented the combined results of the 15 head-to-head comparisons in this meta-analysis.

**Figure 2.**
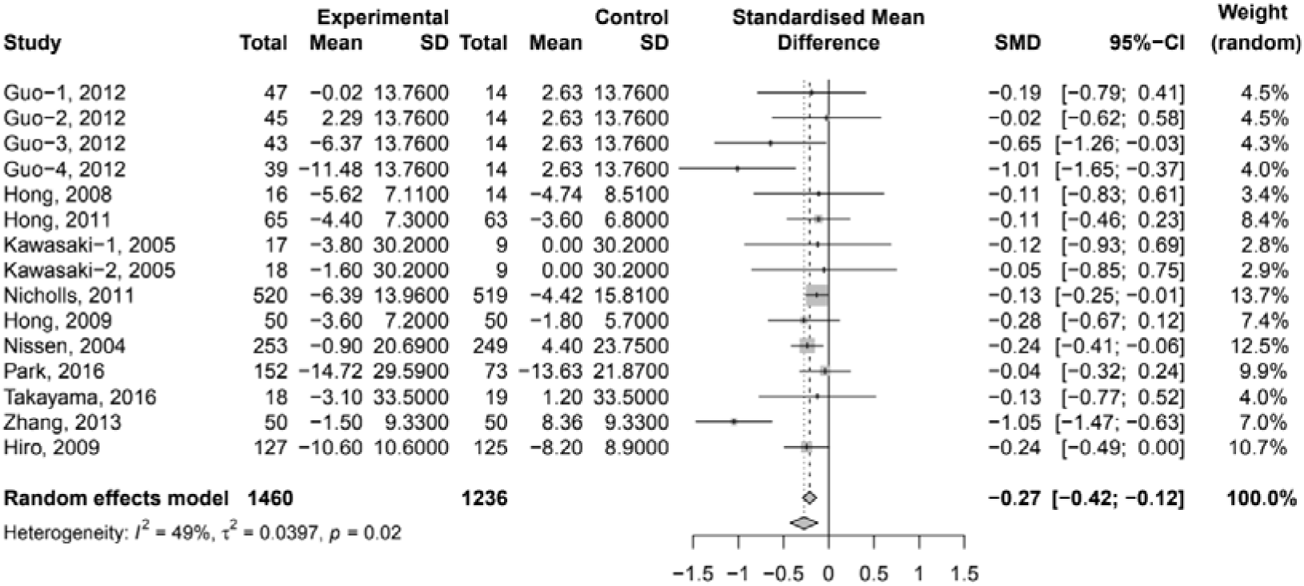
Forest plot of change of TAV/PV. A meta-analysis of 15 statin-treated arms reported a significant reduction in change of TAV/PV (standardized mean difference [SMD]: −0.27, 95% confidence intervals [CI]: −0.42, −0.12), compared with the control arms.

### Effect of statin therapy on change of change of PAV

7 studies (n=2295) reported change of PAV. Heterogeneity test of data from 7 studies shown moderate heterogeneity (*Q*=10.19, *df*=6, *p*=0.12, *I^2^*=41.1%) and random effect model was adopted. Compared with those in the control group, this meta-analysis indicated that patients in the intervention group have a significant reduction in change of PAV (SMD: −0.16, 95% CI: −0.29, −0.03, *p*<0.05). Figure 3 presented the combined results of 7 studies in this meta-analysis.

**Figure 3.**
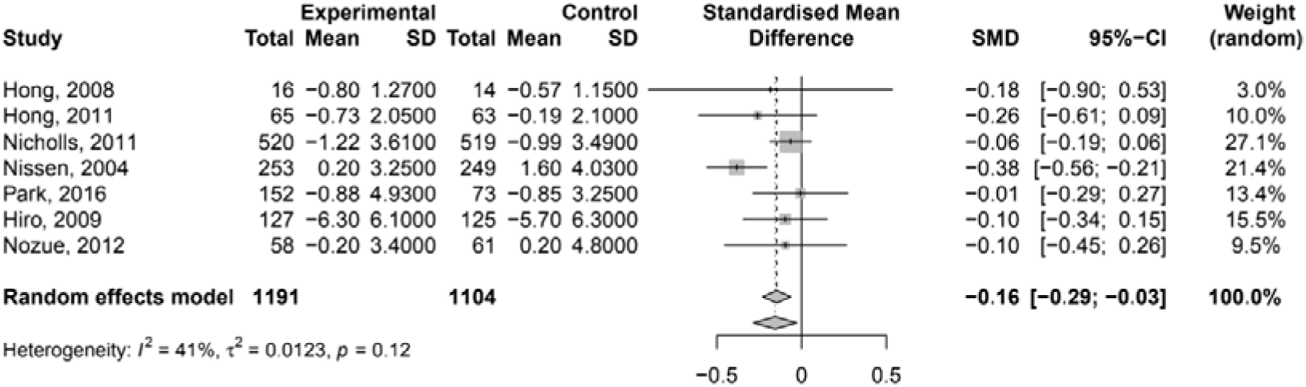
Forest plot of change of PAV A meta-analysis of 7 sstudies reported a significant reduction in change of PAV (standardized mean difference [SMD]: −0.16, 95% confidence intervals [CI]: −0.29, −0.03), compared with the control.

### Meta-regression for SMD in change of TAV/PV

Meta-regression was then employed to test whether the percent change of CRP/hsCRP was associated with the change of TAV/PV. The results of the meta-regression analysis are given in table 2. Model 1 demonstrates that the impact of percent change of CRP/hsCRP on change of TAV/PV was statistically significant (*p*<0.05). The regression coefficient of this independent variable was β=0.0064. Model 2 analyzed the influence of percent change of LDL-C on change of TAV/PV. The results showed that percent change of LDL-C had no significant effect on change of TAV/PV (*p*>0.05). Model 3 incorporates percent changes of CRP/hsCRP and LDL-C. Only percent change of CRP/hsCRP was associated with change of TAV/PV (β=0.0119, *p*<0.05). In Model 4, we entered percent change of CRP/hsCRP, percent change of LDL-C, age, and gender. Among them, only percent change of CRP/hsCRP statistically influences the dependent variable.

### Meta-regression for SMD in change of PAV

Similarly, we performed another meta-regression to explore how the percent change of CRP/hsCRP affects change of PAV. Model 1 used the percent change of CRP/hsCRP as an independent variable. The results indicated that the percent change of CRP/hsCRP (β=0.0086) affects PAV change (*p*<0.01). When the percent change of CRP/hsCRP was higher, change of PAV was greater. Model 2 shows that the percent change of LDL-C was not significantly associated with PAV change (*p*>0.05). In Model 3 (both percent change of CRP/hsCRP and percent change of LDL-C were included as independent variables) and Model 4 (independent variables including percent change of CRP/hsCRP, percent change of LDL-C, age, and gender), multivariable meta-regression analyses did not reveal any significant between independent variables and the change of PAV.

### Publication bias and Sensitivity analysis

Although Begg’s rank correlation (*p*=0.7290) and Egger’s linear regression (*p*=0.2323) tests were not significant, the funnel plot was asymmetric, implying potential publication bias in reporting the effect of statin therapy on change of TAV/PV. Regarding the impact of statin therapy on change of PAV, the number of studies was insufficient to conduct Begg’s test and Egger’s tests. However, the funnel plot also indicated potential publication bias. Funnel plots were presented in Supplementary 4.

Sensitivity analysis by excluding one study each time confirmed that the pooled estimate was consistent among studies with balanced weight. Additional sensitivity analyses were presented in Supplementary 5.

## Discussion

This meta-analysis comprised RCTs using IVUS to measure coronary plaque volume and reporting results of TAV, PAV, or PV changes. Quantitative synthesis revealed a decrease in TAV/PV and PAV levels after statin treatment compared with control. In addition, all studies included in the meta-analysis were RCTs, further confirming that statins are effective drugs for reducing the volume of atherosclerotic plaque in coronary arteries. In addition, our meta-regressions showed that the percent change of CRP/hsCRP reduction was associated with a significant reduction in change of TAV/PV after statin therapy. To the best of our knowledge, this study firstly investigated the association between CRP/hsCRP change and atherosclerotic plaque reduction using the analyses of meta-regressions.

Statins are HMG-COA reductase inhibitors. They reduce CHD incidence due to their lipid-regulating and extra-lipid-regulating effects and are important drugs for the primary and secondary prevention of CHD [25, 26]. The benefits of statins have been demonstrated to be based on stabilization and/or reversal of atherosclerotic plaque [27–30]. Particularly since the introduction of IVUS technology, numerous studies have used it as an important tool for studying coronary plaque. IVUS has recently become the main tool to study the effects of statins on coronary atherosclerotic plaque, and the data obtained by IVUS (such as total plaque volume, plaque cross-sectional area, etc.) served as the primary endpoint in several studies [31, 32].

Recent studies suggest that LDL has been shown to accumulate abnormally in the vascular wall due to endothelial cell dysfunction. Besides, LDL can be converted into ox-LDL, eventually promoting plaque progression [33]. This implies that LDL change is a potential factor affecting plaque volume progression. As a result, we separately included percent change of LDL-C as an independent variable to establish a simple linear regression model, and the results showed that LDL-C change did not influence the result. Moreover, when the percent change of CRP/hsCRP, percent change of LDL-C, age, and gender were simultaneously taken as independent variables to establish the regression model, only the percent change of CRP/hsCRP had a significant impact on TAV/PV. These results indicated that in the included RCTs studies using statins as intervention drugs, the ability of statins to reduce TAV/PV depends on their effect of reducing CRP/hsCRP. The greater the reduction in CRP/hsCRP from baseline after statin treatment, the greater the reduction in TAV/PV. Furthermore, the effect of statins on TAV/PV reduction might not affected by their lipid-regulating effect, nor by age or gender. Previous studies have disclosed that various factors influence the degree of plaque regression under statin therapy. For instance, the statin drug type [34], plaque composition [35], and patient’s age and gender [36]. In addition, clinical trials using IVUS demonstrated a linear relationship between LDL-C levels and reductions in atheroma burden under statin treatment [37]. Despite the well-established causal role of LDL-C in the pathogenesis of atherosclerosis, our findings do not support a reduction in TAV/PV relying on LDL-C levels. Recent investigations have demonstrated that changes in LDL-C levels are unrelated to plaque progression/regression following ezetimibe treatment [38]. This is consistent with our research conclusions. Additionally, previous research has demonstrated that anti-inflammatory therapy alone is beneficial for plaque regression [39]. Considering the pleiotropic nature of statins, CRP/hsCRP is an important indicator of the anti-inflammatory effect of statins. Our findings imply that statins promote plaque regression due to their anti-inflammatory ability, independently of their ability to regulate LDL-C.

However, the percent change of CRP/hsCRP was not significantly associated with PAV change after adjusting for the percent change of LDL-C, age and gender. This could be because only seven trials were included in the regression analysis. The instability of research outcomes is caused by insufficient research data and an excessive number of independent variables.

At present, the main mechanisms of plaque formation include vascular endothelial dysfunction, intimal hyperplasia, lipid accumulation, and inflammatory response. Numerous clinical trials have established that the advantages of statins are based on their pleiotropic properties, such as reducing inflammation, stabilizing plaque, improving vascular endothelial function, suppressing vascular smooth muscle proliferation, and so on [40]. Both basic studies and clinical trials confirmed that the anti-inflammatory ability of statins was beneficial to the prognosis of coronary plaque volume. Basic research shows that atherosclerotic plaques express C-reactive protein (CRP), induce macrophage activation. CRP and type oxidized low-density lipoprotein cholesterol (oxLDL-C) after being converted into foam cells stimulate tissue factor before thrombus formation, endothelial cell expression of adhesion molecules, and vascular endothelial dysfunction, all of which contribute to unstable atherosclerotic plaque. Simultaneously, the expression and release of inflammatory factors are regulated to accelerate atherosclerotic plaque formation [41, 42]. Statins block CRP production and inhibit its pro-inflammatory effects, whereas macrophages in atherosclerotic plaques produce oxygen free radicals that prevent blood vessel inflammation [43]. In previous studies, despite the large body of evidence associating CRP with atherosclerotic lesions, the lack of a direct correlation between its concentration and the extension of atherosclerosis as determined by imaging techniques [4]. Our study confirms that the anti-inflammatory effects of statins have a positive effect on atherosclerotic plaque regression as measured by the IVUS technique. This result suggests that CRP/hsCRP may be a potential therapeutic target in the process of atherosclerosis during statin therapy. Therefore, future research should continue to further study the effect of statin therapy on anti-inflammatory, including reducing serum CRP/hsCRP level.

This study also has some limitations. First of all, we only searched 3 databases. It is possible that some studies in other databases and gray literature are overlooked. Second, although the studies included in this meta-analysis are all RCTs and the quality of evidence is relatively higher, not all studies were double-blind trials. It is possible that performance bias is introduced. The meta-regression analysis (change of PAV as the dependent variable) was performed with 7 trials, which might lead to insufficient statistical power.

In conclusion, our mete-analysis indicated that statins could significantly reduce plaque load measured by TAV/PV and PAV. Further meta-regression revealed that the percent change of CRP/hsCRP was significantly associated with the reduction in plaque volume. The percent change of LDL-C was not significantly associated with TAV/PV change or PAV change. Our results support that statins promote plaque regression through their anti-inflammatory ability and that their ability to reduce plaque volume is unaffected by their ability to reduce LDL-C. This finding will provide new avenues for future research on plaque regression.

## Author contributions

Hua, Ma and Xie conceived and designed the study. Hua, Dina, Ma, Li and Wu performed the statistical analysis. Gao, Hua and Xie drafted and revised the manuscript. Xie and Ma is responsible for the integrity of the work as a whole.

## Data Availability

All data produced in the present study are available upon reasonable request to the authors.

## References

1. McLaren, JE, Michael, DR, Ashlin, TG and Ramji, DP, Cytokines, macrophage lipid metabolism and foam cells: implications for cardiovascular disease therapy, Prog Lipid Res, 50(2011)331–347.https://doi.org/10.1016/j.plipres.2011.04.002

2. Masson, W, Lobo, M, Siniawski, D, Molinero, G, Masson, G, et al., Role of non-statin lipid-lowering therapy in coronary atherosclerosis regression: a meta-analysis and meta-regression, Lipids in Health and Disease, 19(2020).https://doi.org/10.1186/s12944-020-01297-5

3. Ngo-Metzger, Q, Zuvekas, S, Shafer, P, Tracer, H, Borsky, AE, et al., Statin Use in the U.S. for Secondary Prevention of Cardiovascular Disease Remains Suboptimal, J Am Board Fam Med, 32(2019)807–817.https://doi.org/10.3122/jabfm.2019.06.180313

4. Salazar, J, Martínez, MS, Chávez, M, Toledo, A, Añez, R, et al., C-reactive protein: clinical and epidemiological perspectives, Cardiol Res Pract, 2014(2014)605810.https://doi.org/10.1155/2014/605810

5. Sukegawa, H, Maekawa, Y, Yuasa, S, Anzai, A, Kodaira, M, et al., Intensive statin therapy stabilizes C-reactive protein, but not chemokine in stable coronary artery disease treated with an everolimus-eluting stent, Coron Artery Dis, 27(2016)405–411.https://doi.org/10.1097/mca.0000000000000375

6. Ridker, PM, Danielson, E, Fonseca, FA, Genest, J, Gotto, AM, Jr., et al., Rosuvastatin to prevent vascular events in men and women with elevated C-reactive protein, N Engl J Med, 359(2008)2195–2207.https://doi.org/10.1056/NEJMoa0807646

7. Ridker, PM, Everett, BM, Thuren, T, MacFadyen, JG, Chang, WH, et al., Antiinflammatory Therapy with Canakinumab for Atherosclerotic Disease, N Engl J Med, 377(2017)1119–1131.https://doi.org/10.1056/NEJMoa1707914

8. Moher, D, Cook, DJ, Eastwood, S, Olkin, I, Rennie, D, et al., Improving the quality of reports of meta-analyses of randomised controlled trials: the QUOROM statement. Quality of Reporting of Meta-analyses, Lancet, 354(1999)1896–1900.https://doi.org/10.1016/s0140-6736(99)04149-5

9. Moher, D, Liberati, A, Tetzlaff, J and Altman, DG, Preferred reporting items for systematic reviews and meta-analyses: the PRISMA statement, Ann Intern Med, 151(2009)264–269, w264.https://doi.org/10.7326/0003-4819-151-4-200908180-00135

10. Guo, S, Wang, R, Yang, Z, Li, K and Wang, Q, Effects of atorvastatin on serum lipids, serum inflammation and plaque morphology in patients with stable atherosclerotic plaques, Exp Ther Med, 4(2012)1069–1074.https://doi.org/10.3892/etm.2012.722

11. Kawasaki, M, Sano, K, Okubo, M, Yokoyama, H, Ito, Y, et al., Volumetric quantitative analysis of tissue characteristics of coronary plaques after statin therapy using three-dimensional integrated backscatter intravascular ultrasound, J Am Coll Cardiol, 45(2005)1946–1953.https://doi.org/10.1016/j.jacc.2004.09.081

12. Takayama, T, Komatsu, S, Ueda, Y, Fukushima, S, Hiro, T, et al., Comparison of the Effect of Rosuvastatin 2.5 mg vs 20 mg on Coronary Plaque Determined by Angioscopy and Intravascular Ultrasound in Japanese With Stable Angina Pectoris (from the Aggressive Lipid-Lowering Treatment Approach Using Intensive Rosuvastatin for Vulnerable Coronary Artery Plaque [ALTAIR] Randomized Trial), Am J Cardiol, 117(2016)1206–1212.https://doi.org/10.1016/j.amjcard.2016.01.013

13. Zhang, X, Wang, H, Liu, S, Gong, P, Lin, J, et al., Intensive-dose atorvastatin regimen halts progression of atherosclerotic plaques in new-onset unstable angina with borderline vulnerable plaque lesions, J Cardiovasc Pharmacol Ther, 18(2013)119–125.https://doi.org/10.1177/1074248412465792

14. Park, SJ, Kang, SJ, Ahn, JM, Chang, M, Yun, SC, et al., Effect of Statin Treatment on Modifying Plaque Composition: A Double-Blind, Randomized Study, J Am Coll Cardiol, 67(2016)1772–1783.https://doi.org/10.1016/j.jacc.2016.02.014

15. Okazaki, S, Yokoyama, T, Miyauchi, K, Shimada, K, Kurata, T, et al., Early statin treatment in patients with acute coronary syndrome: demonstration of the beneficial effect on atherosclerotic lesions by serial volumetric intravascular ultrasound analysis during half a year after coronary event: the ESTABLISH Study, Circulation, 110(2004)1061–1068.https://doi.org/10.1161/01.Cir.0000140261.58966.A4

16. Lee, SW, Hau, WK, Kong, SL, Chan, KK, Chan, PH, et al., Virtual histology findings and effects of varying doses of atorvastatin on coronary plaque volume and composition in statin-naive patients: the VENUS study, Circ J, 76(2012)2662–2672.https://doi.org/10.1253/circj.cj-12-0325

17. Matsushita, K, Hibi, K, Komura, N, Akiyama, E, Maejima, N, et al., Effects of 4 Statins on Regression of Coronary Plaque in Acute Coronary Syndrome, Circ J, 80(2016)1634–1643.https://doi.org/10.1253/circj.CJ-15-1379

18. Hiro, T, Kimura, T, Morimoto, T, Miyauchi, K, Nakagawa, Y, et al., Effect of intensive statin therapy on regression of coronary atherosclerosis in patients with acute coronary syndrome: a multicenter randomized trial evaluated by volumetric intravascular ultrasound using pitavastatin versus atorvastatin (JAPAN-ACS [Japan assessment of pitavastatin and atorvastatin in acute coronary syndrome] study), J Am Coll Cardiol, 54(2009)293–302.https://doi.org/10.1016/j.jacc.2009.04.033

19. Hong, MK, Park, DW, Lee, CW, Lee, SW, Kim, YH, et al., Effects of statin treatments on coronary plaques assessed by volumetric virtual histology intravascular ultrasound analysis, JACC Cardiovasc Interv, 2(2009)679–688.https://doi.org/10.1016/j.jcin.2009.03.015

20. Hong, YJ, Jeong, MH, Chung, JW, Sim, DS, Cho, JS, et al., The effects of rosuvastatin on plaque regression in patients who have a mild to moderate degree of coronary stenosis with vulnerable plaque, Korean circulation journal, 38(2008)366–373.https://doi.org/10.4070/kcj.2008.38.7.366

21. Hong, YJ, Jeong, MH, Hachinohe, D, Ahmed, K, Choi, YH, et al., Comparison of effects of rosuvastatin and atorvastatin on plaque regression in Korean patients with untreated intermediate coronary stenosis, Circ J, 75(2011)398–406.https://doi.org/10.1253/circj.cj-10-0658

22. Nicholls, SJ, Ballantyne, CM, Barter, PJ, Chapman, MJ, Erbel, RM, et al., Effect of two intensive statin regimens on progression of coronary disease, N Engl J Med, 365(2011)2078–2087.https://doi.org/10.1056/NEJMoa1110874

23. Nissen, SE, Tuzcu, EM, Schoenhagen, P, Brown, BG, Ganz, P, et al., Effect of intensive compared with moderate lipid-lowering therapy on progression of coronary atherosclerosis: a randomized controlled trial, Jama, 291(2004)1071–1080.https://doi.org/10.1001/jama.291.9.1071

24. Nozue, T, Yamamoto, S, Tohyama, S, Umezawa, S, Kunishima, T, et al., Statin treatment for coronary artery plaque composition based on intravascular ultrasound radiofrequency data analysis, Am Heart J, 163(2012)191–199.e191.https://doi.org/10.1016/j.ahj.2011.11.004

25. Group, LS, Long-term effectiveness and safety of pravastatin in 9014 patients with coronary heart disease and average cholesterol concentrations: the LIPID trial follow-up, Lancet, 359(2002)1379–1387.https://doi.org/10.1016/S0140-6736(02)08351-4

26. Pedersen, TR, Kjekshus, J, Berg, K, Haghfelt, T, Faergeman, O, et al., Randomised trial of cholesterol lowering in 4444 patients with coronary heart disease: the Scandinavian Simvastatin Survival Study (4S). 1994, Atheroscler Suppl, 5(2004)81–87.https://doi.org/10.1016/j.atherosclerosissup.2004.08.027

27. Böse, D, von Birgelen, C and Erbel, R, Intravascular ultrasound for the evaluation of therapies targeting coronary atherosclerosis, J Am Coll Cardiol, 49(2007)925–932.https://doi.org/10.1016/j.jacc.2006.08.067

28. Endo, H, Dohi, T, Miyauchi, K, Kuramitsu, S, Kato, Y, et al., Clinical significance of non-culprit plaque regression following acute coronary syndrome: A serial intravascular ultrasound study, J Cardiol, 74(2019)102–108.https://doi.org/10.1016/j.jjcc.2018.12.023

29. Jensen, LO, Thayssen, P, Pedersen, KE, Stender, S and Haghfelt, T, Regression of coronary atherosclerosis by simvastatin: a serial intravascular ultrasound study, Circulation, 110(2004)265–270.https://doi.org/10.1161/01.Cir.0000135215.75876.41

30. Yamada, T, Azuma, A, Sasaki, S, Sawada, T and Matsubara, H, Randomized evaluation of atorvastatin in patients with coronary heart disease: a serial intravascular ultrasound study, Circ J, 71(2007)1845–1850.https://doi.org/10.1253/circj.71.1845

31. Banach, M, Serban, C, Sahebkar, A, Mikhailidis, DP, Ursoniu, S, et al., Impact of statin therapy on coronary plaque composition: a systematic review and meta-analysis of virtual histology intravascular ultrasound studies, BMC Med, 13(2015)229.https://doi.org/10.1186/s12916-015-0459-4

32. D’Ascenzo, F, Agostoni, P, Abbate, A, Castagno, D, Lipinski, MJ, et al., Atherosclerotic coronary plaque regression and the risk of adverse cardiovascular events: a meta-regression of randomized clinical trials, Atherosclerosis, 226(2013)178–185.https://doi.org/10.1016/j.atherosclerosis.2012.10.065

33. Barua, RS, Ambrose, JA, Srivastava, S, DeVoe, MC and Eales-Reynolds, LJ, Reactive oxygen species are involved in smoking-induced dysfunction of nitric oxide biosynthesis and upregulation of endothelial nitric oxide synthase: an in vitro demonstration in human coronary artery endothelial cells, Circulation, 107(2003)2342–2347.https://doi.org/10.1161/01.Cir.0000066691.52789.Be

34. Qian, C, Wei, B, Ding, J, Wu, H, Cai, X, et al., Meta-analysis comparing the effects of rosuvastatin versus atorvastatin on regression of coronary atherosclerotic plaques, Am J Cardiol, 116(2015)1521–1526.https://doi.org/10.1016/j.amjcard.2015.08.010

35. Kwon, O, Kang, SJ, Kang, SH, Lee, PH, Yun, SC, et al., Relationship Between Serum Inflammatory Marker Levels and the Dynamic Changes in Coronary Plaque Characteristics After Statin Therapy, Circ Cardiovasc Imaging, 10(2017).https://doi.org/10.1161/circimaging.116.005934

36. Dai, J, Hou, J, Xing, L, Jia, H, Hu, S, et al., Is age an important factor for vascular response to statin therapy? A serial optical coherence tomography and intravascular ultrasound study, Coron Artery Dis, 28(2017)209–217.https://doi.org/10.1097/mca.0000000000000465

37. Nicholls, SJ, Puri, R, Anderson, T, Ballantyne, CM, Cho, L, et al., Effect of Evolocumab on Progression of Coronary Disease in Statin-Treated Patients: The GLAGOV Randomized Clinical Trial, Jama, 316(2016)2373–2384.https://doi.org/10.1001/jama.2016.16951

38. Spence, JD and Solo, K, Resistant Atherosclerosis: The Need for Monitoring of Plaque Burden, Stroke, 48(2017)1624–1629.https://doi.org/10.1161/strokeaha.117.017392

39. Ridker, PM, Howard, CP, Walter, V, Everett, B, Libby, P, et al., Effects of interleukin-1β inhibition with canakinumab on hemoglobin A1c, lipids, C-reactive protein, interleukin-6, and fibrinogen: a phase IIb randomized, placebo-controlled trial, Circulation, 126(2012)2739–2748.https://doi.org/10.1161/circulationaha.112.122556

40. Dupuis, J, Tardif, JC, Rouleau, JL, Ricci, J, Arnold, M, et al., Intensity of lipid lowering with statins and brachial artery vascular endothelium reactivity after acute coronary syndromes (from the BRAVER trial), Am J Cardiol, 96(2005)1207–1213.https://doi.org/10.1016/j.amjcard.2005.06.057

41. Avan, A, Tavakoly Sany, SB, Ghayour-Mobarhan, M, Rahimi, HR, Tajfard, M, et al., Serum C-reactive protein in the prediction of cardiovascular diseases: Overview of the latest clinical studies and public health practice, J Cell Physiol, 233(2018)8508–8525.https://doi.org/10.1002/jcp.26791

42. Cicci, JD, Iyer, P, Clarke, MM and Mazzella, AJ, Aspirin for the Primary Prevention of Cardiovascular Disease: A Review of the Literature and Considerations for Clinical Practice, Cardiol Rev, 28(2020)98–106.https://doi.org/10.1097/crd.0000000000000297

43. Endres, M, Statins: potential new indications in inflammatory conditions, Atheroscler Suppl, 7(2006)31–35.https://doi.org/10.1016/j.atherosclerosissup.2006.01.005

