## Supplementary material for "Effects of statin therapy on coronary plaque volume by decreasing CRP/hsCRP levels: A meta-regression of randomized controlled trials": Tables

**Table1** Main characteristics and findings of included studies.

| First author, published year | Country | Study duration | Therapy  (mg/d) | Participants (n) | Age  (years) | Male  (%) | CRP  /hsCRP | Percent change of CRP/hsCRP (%) | Percent change of LDL-C  (%) | Change of TAV/PV (mm^3^) | Change of PAV  (%) |
| --- | --- | --- | --- | --- | --- | --- | --- | --- | --- | --- | --- |
| Hong, 2008 | Korea | 12 months | Ros 20 | 16 | 60±8 | 75 | hsCRP | −94.35 | −46.38 | −5.62±7.71 | −0.80±1.27 |
|  |  |  | Ato 40 | 14 | 62±90 | 43 |  | −93.85 | −43.31 | −4.74±8.51 | −0.57±1.15 |
| Hong, 2011 | Korea | 11 months | Ros 20 | 65 | 59±10 | 75 | hsCRP | −80 | −49.18 | −4.4±7.3 | −0.73±2.05 |
|  |  |  | Ato 40 | 63 | 58±10 | 73 |  | −89.25 | −40.17 | −3.6±6.8 | −0.19±2.10 |
| Kawasaki, 2005 | Japan | 6 months | Ato 20 | 18 | 66±8.7 | 70.6 | CRP | −65 | −39 | −3.8±32.2 | / |
|  |  |  | Pra 20 | 17 | 67±7.8 | 72.2 |  | −18 | −32 | −1.6±32.1 | / |
|  |  |  | Diet | 17 | 66±6.4 | 82.4 |  | 17 | −2 | 0±29.9 | / |
| Nicholls, 2011 | American, et al | 104 weeks | Ros 40 | 520 | 57.4±8.6 | 72.9 | CRP | −35.29 | −47.83 | −6.39±13.96 | −1.22±3.61 |
|  |  |  | Ato 80 | 519 | 57.9±8.5 | 74.4 |  | −33.33 | −41.45 | −4.42±15.81 | −0.99±3.49 |
| Nissen, 2004 | American | 18 months | Ato 80 | 253 | 55.8 ±9.8 | 71 | CRP | −36.4 | −46.3 | −0.9±20.69 | 0.2±3.25 |
|  |  |  | Pra 40 | 249 | 56.6±9.2 | 73 |  | −5.2 | −25.2 | 4.4±23.75 | 1.6±4.03 |
| Nozue, 2012 | Japan | 8 months | Pit 4 | 58 | 66±9 | 90 | hsCRP | −75 | −41 | / | −0.2±3.4 |
|  |  |  | Pra 20 | 61 | 67±11 | 77 |  | −75 | −29 | / | 0.2±4.8 |
| Park,  2016 | Korea | 12 months | Ros 40 | 152 | 62.6±9.3 | 71 | hsCRP | −52.38 | −43.87 | −14.72±29.59 | −0.88±4.93 |
|  |  |  | Ros 10 | 73 | 61.8±8.9 | 77 |  | −47.83 | −27.90 | −13.63±21.87 | −0.85±3.25 |
| Takayama, 2016 | Japan | 48 weeks | Ros 20 | 18 | 65.1±10.1 | 72 | hsCRP | −65 | −50 | −3.1±33.5 | / |
|  |  |  | Ros 2.5 | 19 | 63.8±8.5 | 83 |  | −60 | −30 | 1.2±33.5 | / |
| Hiro,  2009 | Japan | 8-12 months | Ato 20 | 127 | 62.4±10.6 | 81.1 | hsCRP | −95.4 | −35.8 | −10.6±10.6 | −6.3±6.1 |
|  |  |  | Pit 4 | 125 | 62.5±11.5 | 82.4 |  | −97.3 | −36.2 | −8.2±8.9 | −5.7±6.3 |
| Hong, 2009 | Korea | 12 months | Ros 10 | 50 | 59±9 | 74 | CRP | −57.14 | −44.83 | −3.6±7.2 | / |
|  |  |  | Sim 20 | 50 | 58±10 | 80 |  | −29.41 | −34.45 | −1.8±5.7 | / |
| Zhang, 2013 | China | 9 months | Ato 80 | 50 | 64.5±13.8 | 62 | hsCRP | −66.36 | −40.91 | −1.5±9.33 | / |
|  |  |  | Ato 20 | 50 | 65.5±6.2 | 58 |  | −37.41 | −24.58 | 8.36±9.33 | / |
| Guo,  2012 | China | 6 months | Ato 10 | 47 | 62.64±12.00 | 85.1 | hsCRP | 11.59 | −22.11 | −0.02±13.76 | / |
|  |  |  | Ato 20 | 45 | 59.18±8.48 | 80.0 |  | 0.39 | −31.16 | 2.29±13.76 | / |
|  |  |  | Ato 40 | 43 | 58.91±12.90 | 95.3 |  | −13.94 | −36.21 | −6.37±13.76 | / |
|  |  |  | Ato 80 | 39 | 58.95±9.68 | 87.2 |  | −41.15 | −36.04 | −11.48±13.76 | / |
|  |  |  | Placebo | 54 | 62.07±8.51 | 88.9 |  | 35.50 | 1.02 | 2.63±13.76 | / |

^a^ Ros: rosuvastatin; Ato: atorvastatin; Pra: pravastatin; Pit: pitavastatin; Sim: simvastatin.

**Table 2** meta-regression analysis for SMD in change of TAV/PV

|  | Model 1 | Model 2 | Model 3 | Model 4 |
| --- | --- | --- | --- | --- |
| Variables | β | β | β | β |
| Intercept | −0.1463 | −0.1513 | **−0.2419*** | 0.768 |
| Percent change of CRP/hsCRP | **0.0064*** | —— | **0.0119*** | **0.0124**** |
| Percent change of LDL-C | —— | 0.0075 | −0.0129 | −0.0137 |
| Age | —— | —— | —— | −0.0248 |
| Gender | —— | —— | —— | 0.0063 |

^a^ **p*<0.05; and ***p*<0.01

**Table 3** meta-regression analysis for SMD in change of PAV

|  | Model 1 | Model 2 | Model 3 | Model 4 |
| --- | --- | --- | --- | --- |
| Variables | β | β | β | β |
| Intercept | −0.0833 | −0.0217 | −0.0735 | −0.6353 |
| Percent change of CRP/hsCRP | **0.0086**** | —— | 0.0079 | 0.0045 |
| Percent change of LDL-C | —— | 0.0127 | 0.0015 | 0.0038 |
| Age | —— | —— | —— | 0.0158 |
| Gender | —— | —— | —— | −0.0052 |

^a^ **p*<0.05; and ***p*<0.01
