## Supplementary 1-5 for "Effects of statin therapy on coronary plaque volume by decreasing CRP/hsCRP levels: A meta-regression of randomized controlled trials"

**Supplementary 1. Details of search syntax**

| **Database** | **Search Syntax** |
| --- | --- |
| **Pubmed** | ((statin[Title/Abstract]) OR (hydroxy-methyl-glutaryl-CoA[Title/Abstract]) OR (HMG-COA[Title/Abstract]) OR (pravastatin[Title/Abstract]) OR (lovastatin[Title/Abstract]) OR (simvastatin[Title/Abstract]) OR (Atorvastatin[Title/Abstract]) OR (fluvastatin[Title/Abstract]) OR (Rosuvastatin[Title/Abstract]) OR (Pitavastatin[Title/Abstract]))  AND ((intravascular ultrasound[Title/Abstract]) OR (IVUS [Title/Abstract]) OR (plaque [Title/Abstract]) OR (atheroma[Title/Abstract])) AND ((intravascular ultrasound[Title/Abstract]) OR (IVUS [Title/Abstract]) OR (coronary[Title/Abstract])) AND (Clinical Trial[ptyp]) |
| **EMBASE** | (“statin”:ab,ti OR “hydroxy-methyl-glutaryl-CoA”:ab,ti OR “HMG-COA”:ab,ti OR “pravastatin”:ab,ti OR “lovastatin”:ab,ti OR “simvastatin”:ab,ti OR “Atorvastatin”:ab,ti OR “fluvastatin”:ab,ti OR “Rosuvastatin”:ab,ti OR “Pitavastatin”:ab,ti)  AND (“intravascular ultrasound”:ab,ti OR “IVUS”:ab,ti OR “plaque”:ab,ti OR “atheroma”:ab,ti) AND (“intravascular ultrasound”:ab,ti OR “IVUS”:ab,ti OR “coronary”:ab,ti) AND ([controlled clinical trial]/lim OR [randomized controlled trial]/lim) |
| **Cochrane** | (“statin” OR “hydroxy-methyl-glutaryl-CoA” OR “HMG-COA” OR “pravastatin” OR “lovastatin” OR “simvastatin” OR “Atorvastatin” OR “fluvastatin” OR “Rosuvastatin” OR “Pitavastatin”): ti,ab,kw  AND (“intravascular ultrasound” OR “IVUS” OR “plaque” OR “atheroma”): ti,ab,kw AND (“intravascular ultrasound” OR “IVUS” OR “coronary”): ti,ab,kw in Trials |

**Supplementary 2. Details of data extraction.**

| First author, published year | Therapy | CRP/hs-CRP (mg/L) | |  | LDL-C (mg/dL) | |  | PV/TAV (mm^3^/L) | |  | PAV (%) | |
| --- | --- | --- | --- | --- | --- | --- | --- | --- | --- | --- | --- | --- |
|  |  | Baseline | Follow-up |  | Baseline | Follow-up |  | Baseline | Follow-up |  | Baseline | Follow-up |
| Hong, 2008 | Ros 20 | 12.4±1.9 | 0.7±1.9 |  | 121±45 | 65±25 |  | 252±80 | 246±79 |  | 45.5±6.1 | 44.7±6.0 |
|  | Ato 40 | 13.0±22 | 0.8±1.0 |  | 127±37 | 72±26 |  | 288±98 | 283±98 |  | 46.9±4.0 | 46.3±4.6 |
| Hong, 2011 | Ros 20 | 5.0±11.7 | 1.0±1.4 |  | 122±37 | 62±20 |  | 166±93 | 166±93 |  | 48.0±6.1 | 47.3±6.5 |
|  | Ato 40 | 9.3±20.3 | 1.0±1.8 |  | 117±38 | 70±24 |  | 190±119 | 190±119 |  | 49.9±6.1 | 49.7±6.5 |
| Nicholls, 2011 | Ros 40 | 1.7  (0.8–3.8) | 1.1  (0.5–2.4) |  | 120.0±27.3 | 62.6±1.0 |  | 144.1±60.8 | 135.7±57.7 |  | 36.7±8.2 | 35.4±8.2 |
|  | Ato 80 | 1.5  (0.8–3.3) | 1.0  (0.5–2.0) |  | 119.9±28.9 | 70.2±1.0 |  | 144.2±63.8 | 138.5±63.2 |  | 36.0±8.3 | 34.9±8.1 |
| Nissen, 2004 | Ato 80 | 2.8  (3.0) | 1.8  (3.7) |  | 150.2  (27.9) | 78.9  (30.2) |  | 184.4  (115.7) | 183.9  (108.8) |  | 38.4  (11.27) | 39.0  (10.8) |
|  | Pra 40 | 3.0  (2.9) | 2.9  (3.0) |  | 150.2  (25.9) | 110.4  (25.8) |  | 194.5  (114.8) | 199.6  (112.3) |  | 39.5  (10.77) | 41.4  (10.0) |
| Nozue, 2012 | Pit 4 | 3.76 (1.22-9.22) | 0.731 (0.315-1.78) |  | 126±28 | 74±22 |  | / | / |  | 55.2±6.1 | 55.0±6.0 |
|  | Pra 20 | 4.23 (1.21-9.26) | 0.525 (0.285-1.63) |  | 137±35 | 95±23 |  | / | / |  | 53.9±7.8 | 54.1±7.8 |
| Park, 2016 | Ros 40 | 2.1±3.8 | 1.0±1.5 |  | 105.3±32.8 | 59.1±22.2 |  | 194.7  (155.3 – 234.7) | 177.8  (148.3 – 221.9) |  | 51.8  (46.1 – 57.0) | 50.0  (43.7 – 57.2) |
|  | Ros 10 | 2.3±3.9 | 1.2±1.5 |  | 109.3±40.9 | 78.8±27.8 |  | 187.3 (145.4–244.1) | 164.1  (131.2 – 214.8) |  | 50.5  (43.9 – 55.3) | 49.1  (42.9 – 55.7) |

**Supplementary 2.** (continued )

| First author, published year | Therapy | CRP/hs-CRP (mg/L) | |  | LDL-C (mg/dL) | |  | PV/TAV (mm^3^/L) | |  | PAV (%) | |
| --- | --- | --- | --- | --- | --- | --- | --- | --- | --- | --- | --- | --- |
|  |  | Baseline | Follow-up |  | Baseline | Follow-up |  | Baseline | Follow-up |  | Baseline | Follow-up |
| Takayama, 2016 | Ros 20 | 0.200± 0.163 | 0.07± 0.1 |  | 130.3±25.5 | 61.7±16.5 |  | 56.5±34.2 | 53.4±32.3 |  | / | / |
|  | Ros 2.5 | 0.225± 0.196 | 0.09±0.12 |  | 130.9±28.5 | 89.7±29 |  | 58.1±33.5 | 59.3±31.7 |  | / | / |
| Hiro, 2009 | Ato 20 | 14.9  (4.7–62.4) | 0.53  (0.26–1.2) |  | 133.8±31.4 | 84.1±27.4 |  | 63.9±33.9 | 53.3±31.7 |  | 50.5±9.7 | 44.3±10.7 |
|  | Pit 4 | 19.9  (7.1–71.1) | 0.54  (0.36–1.1) |  | 130.9±33.3 | 81.1±23.4 |  | 49.8±28.8 | 41.6±25.0 |  | 49.4±10.8 | 43.7±11.0 |
| Hong, 2009 | Ros 10 | 2.1±2.0 | 0.9±0.7 |  | 116±28 | 64±21 |  | 91.5±27.5 | 87.8±27.8 |  | / | / |
|  | Sim 20 | 1.7±2.2 | 1.2±1.2 |  | 119±30 | 78±20 |  | 88.3±26.9 | 86.3±26.8 |  | / | / |
| Zhang, 2013 | Ato 80 | 5.38±1.48 | 1.81±0.91 |  | 105.55± 22.65 | 62.37± 15.93 |  | 43.20±6.33 | 41.70±4.60 |  | / | / |
|  | Ato 20 | 5.40±1.38 | 3.38±1.28 |  | 106.13± 20.52 | 80.04± 17.77 |  | 42.32±9.33 | 50.68±9.78 |  | / | / |
| Guo, 2012 | Ato 10 | 6.04±2.52 | 6.74±2.20 |  | 3.03±0.70 | 2.36±0.50 |  | 38.07±13.94 | 38.05±13.56 |  | / | / |
|  | Ato 20 | 5.09±1.94 | 5.07±1.72 |  | 2.92±0.62 | 2.01±0.18 |  | 33.83±10.56 | 36.12±11.96 |  | / | / |
|  | Ato 40 | 5.67±2.22 | 4.88±1.52 |  | 2.90±0.34 | 1.85±0.22 |  | 37.06±12.01 | 30.69±8.12 |  | / | / |
|  | Ato 80 | 6.10±2.12 | 3.59±1.07 |  | 2.83±0.66 | 1.81±0.32 |  | 36.47±14.68 | 24.99±1.01 |  | / | / |
|  | Placebo | 5.07±1.80 | 6.87±2.62 |  | 2.94±0.72 | 2.97±0.63 |  | 34.83±13.76 | 37.46±15.80 |  | / | / |

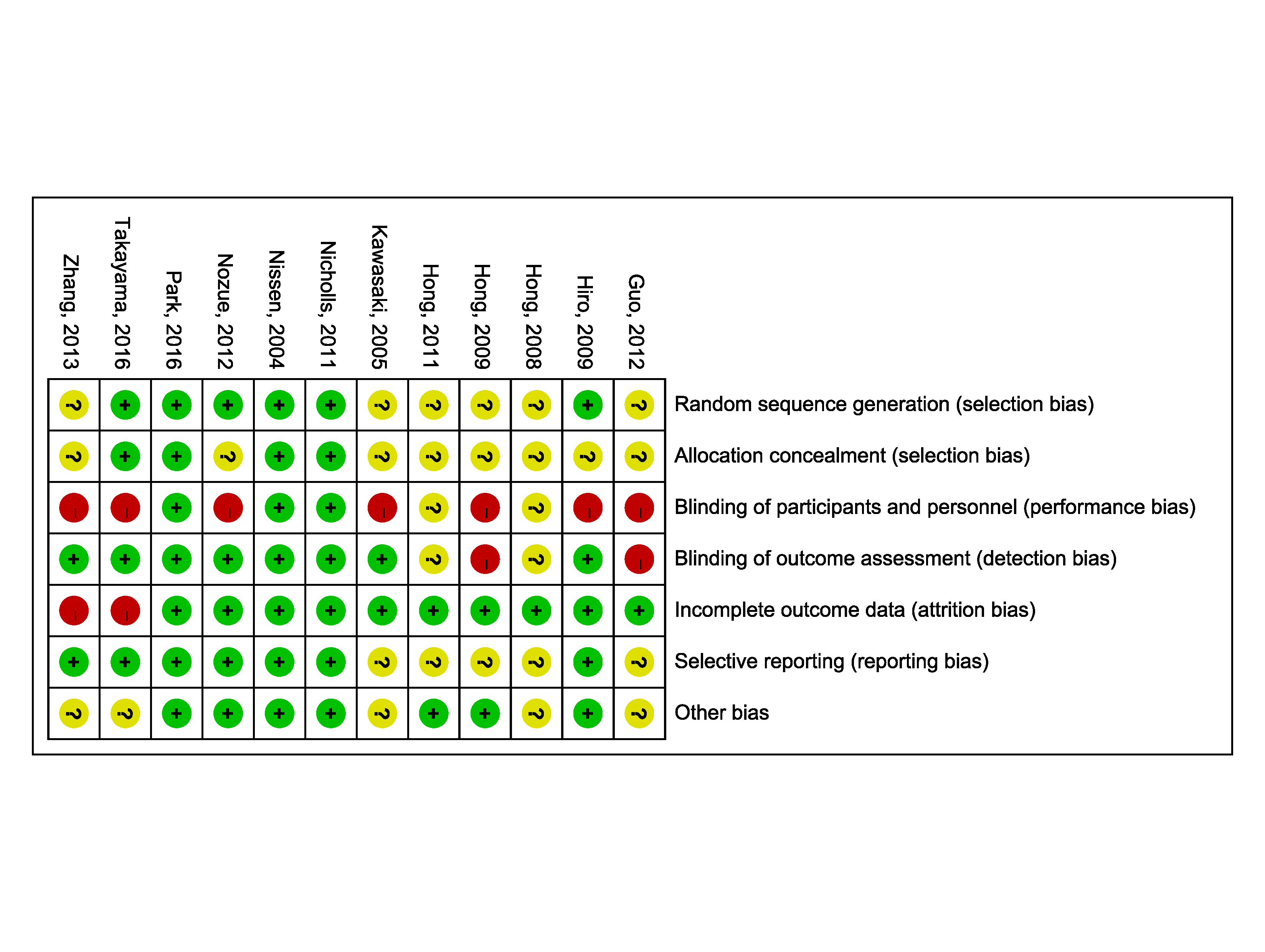

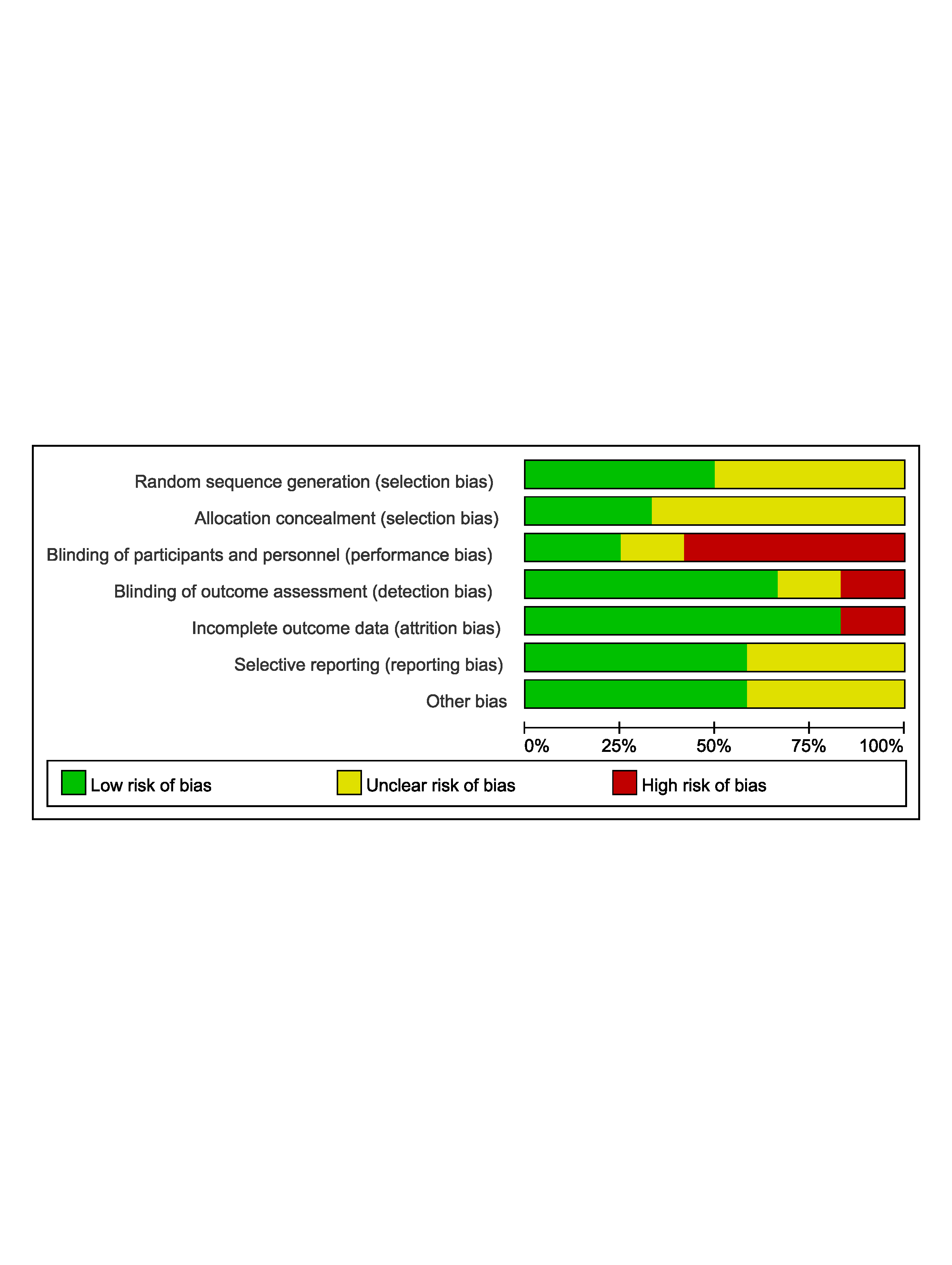

**Supplementary 3** The Cochrane risk of bias assessment

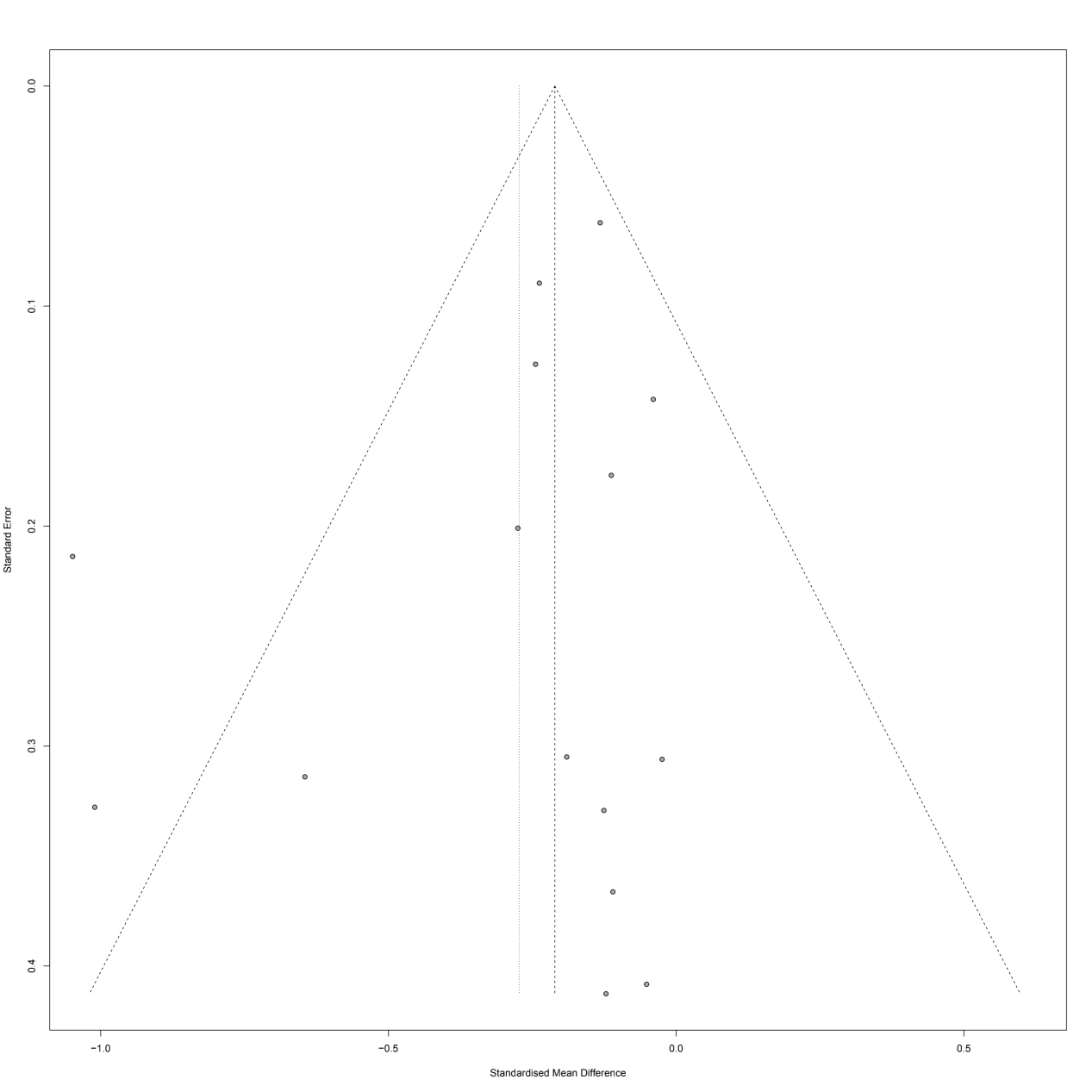

**Supplementary 4a** Funnel plot detailing publication bias in the studies reporting the impact of statin therapy on TAV/PV.

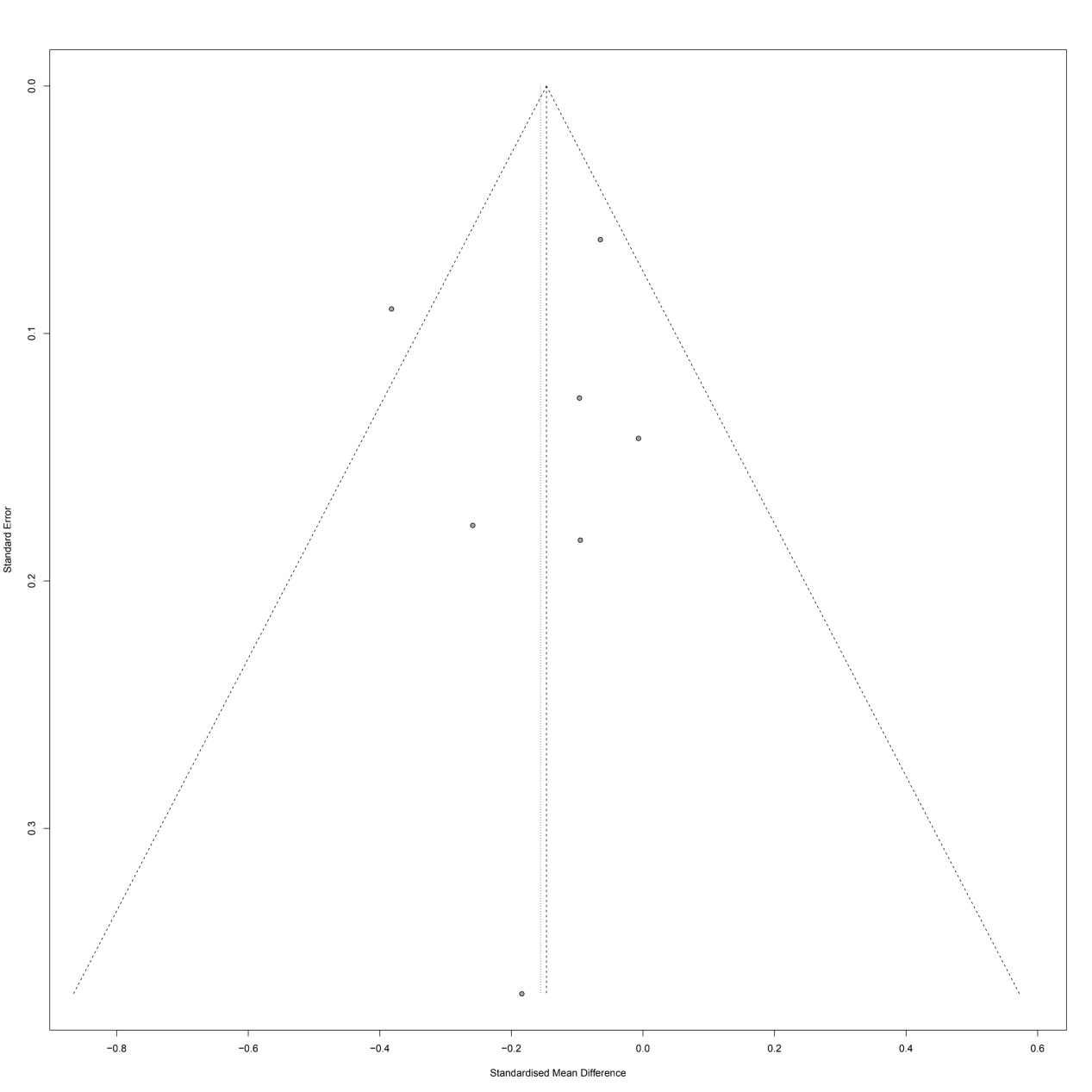

**Supplementary 4b** Funnel plot detailing publication bias in the studies reporting the impact of statin therapy on PAV.

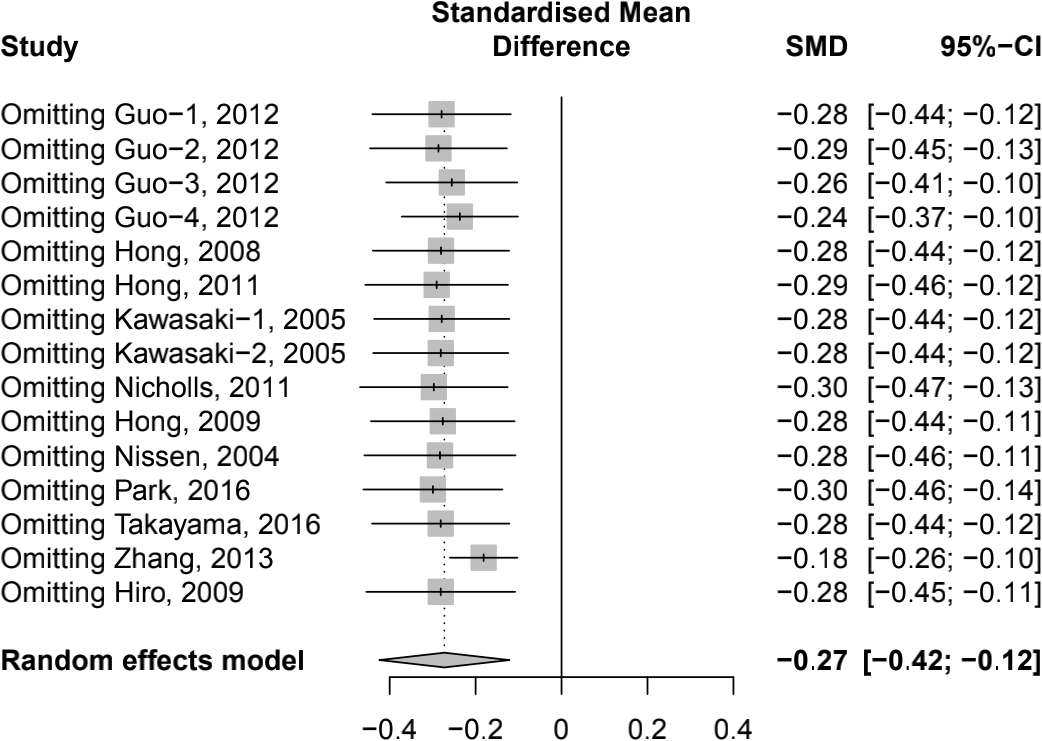

**Appendix 5a** Sensitivity analysis of change of TAV/PV.

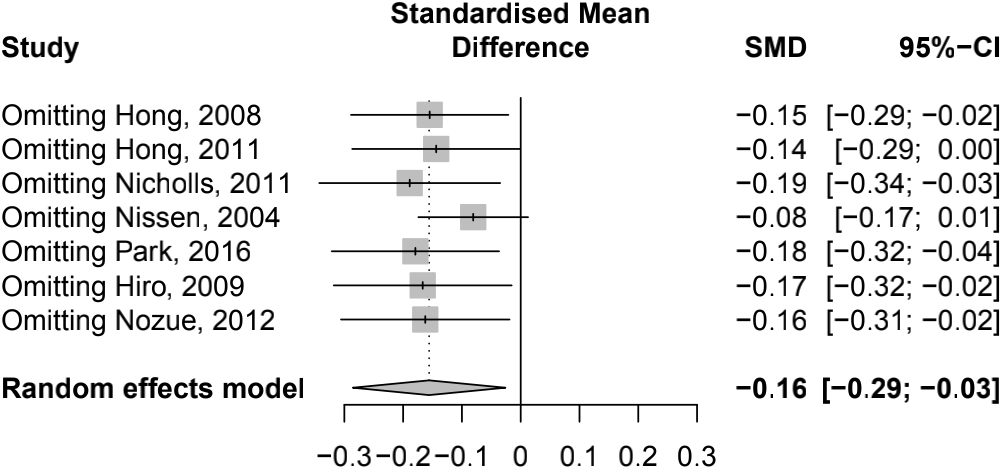

**Supplementary 5a** Sensitivity analysis of change of PAV.
